# Dupilumab-associated ocular surface disease: incidence, management and long-term sequelae

**DOI:** 10.1101/2020.06.16.20124909

**Authors:** Magdalena Z Popiela, Ramez Barbara, Andrew M J Turnbull, Emma Corden, Beatrice Suarez Matinez-Falero, Daniel O’Driscoll, Michael R Ardern-Jones, Parwez N Hossain

## Abstract

**Objective:** To determine the incidence of ocular surface disease in patients with atopic dermatitis (AD) treated with dupilumab at a tertiary, university hospital. To describe the features of dupilumab-associated ocular surface disease, establish the need for treatment and report any long-term effects on the ocular surface.

**Methods:** A retrospective analysis of consecutive patients treated with dupilumab for AD between January 2017 and August 2019 was undertaken. Data was collected on demographics, incidence and type of ocular disease features, natural history and treatment.

**Results:** 50% (14/28) patients developed ocular symptoms with a mean time of onset of 6.75 (+/- 6.1) weeks from starting dupilumab. 69% of these (9/13) were diagnosed with conjunctivitis - associated with cicatrisation in two patients and periorbital skin changes in four. Of these nine, four had prior history of atopic keratoconjunctivitis. All were treated with topical steroids; two required additional ciclosporin drops. 67% (6/9) patients developed chronic ocular inflammation requiring maintenance drops at a mean of 16 (+/- 6.9) months of follow up. All patients had improvement in their AD severity; only one patient discontinued dupilumab due to ocular side effects.

**Conclusion:** The rate of dupilumab-associated ocular surface disease was 32%. Periorbital skin changes and conjunctival cicatrisation were noted in association with conjunctivitis. Ocular surface disease improved on topical steroids and ciclosporin but 67% of patients needed on-going treatment. Patients should be referred to an ophthalmologist prior to starting dupilumab as a large proportion develops chronic ocular inflammation.

## Introduction

Dupilumab is the first biologic approved for use in treatment of moderate to severe atopic dermatitis (AD).^1-5^ Dupilumab targets interleukin (IL)-4 receptor blocking IL-4 and IL-13 signalling pathway. It has also shown promise in treatment of asthma, chronic rhinosinusitis with nasal polyposis and eosinophilic oesophagitis.^1,2^

Dupilumab has been found to significantly improve signs and symptoms of AD and is considered a safe treatment for long-term use.^6^ Conjunctivitis is a known side effect with rates in initial trials varying between 8.6% (CHRONOS) to 28 % (LIBERTY AD CAFÉ). Conjunctivitis is considered mild and self-limiting in the majority of cases, with fewer than 0.5% of patients suffering with a severe form necessitating drug cessation.^2^

Current International Eczema Council recommendations allow for dupilumab to be started in patients with prior ocular surface disease and to continue dupilumab in the event of conjunctivitis occurring.^7^ Prior history of ocular surface disease and more severe AD are considered risk factors for the development of dupilumab-associated conjunctivitis.^8,9^

Since the start of clinical use, apart from self-limiting conjunctivitis other ocular features have been described; severe follicular conjunctivitis ^10,11^, limbal nodules ^4,10^, blepharo-conjunctivitis ^10-13^, cicatricial ectropion ^14,15^, keratitis ^10^ and dry eyes ^16^. This has led to the use of the term dupilumab-associated ocular surface disease to encompass various clinical presentations.^8,9,15^ A much higher incidence of ocular surface problems has been reported in clinical practice than in the initial drug trials - as high as 70% in one series.^4^ No data is available on the longer-term effects of dupilumab on the ocular surface. Our study aimed to establish local incidence of dupilumab-associated ocular surface disease, describe its features, establish the need for treatment and identify any long-term sequelae.

## Methods

The Health Research Authority and Health and Care Research Wales (HCRW) approval was granted in November 2019 (19/HRA/5882) for a retrospective case review of all patients developing dupilumab-associated eye disease at Southampton General Hospital-a large tertiary referral centre in the United Kingdom.

Patients prescribed dupilumab between January 2017 and August 2019 were identified from the severe atopic dermatitis clinic in the dermatology department. Their details were checked in the electronic patient records to identify those who attended ophthalmology clinics. Data for patients who developed eye symptoms was collected retrospectively in February 2020 to allow for at least six months of ophthalmic follow up. Presenting ocular symptoms, signs on initial review, treatment, visual acuity at first and last follow up and any side effects of topical therapy were noted. Long-term use of treatment was recorded. Eczema Severity and Index scores (EASI) were recorded at pre-treatment and at 16 weeks post dupilumab initiation, as well as the percentage of patients reaching 50 % and 75 % improvement in EASI at 16 weeks (EASI 50 and EASI 75 respectively).

### Statistics

Because of only a few studies for reference, the study is classed as an exploratory research study. Data was analysed using descriptive statistics using The SAMPL Guidelines as reference (https://www.equator-network.org/wp-content/uploads/2013/03/SAMPL-Guidelines-3-13-13.pdf) with the statistical tools within Microsoft Excel 2014 (Microsoft Corp., Redmond, WA). Logistic regression and Mann-Whitney U test were used to test for differences in demographic data between patients with and without ocular symptoms. Statistical significance was defined with a P value of equal to or less than 0.05. All statistical analyses were performed using SPSS software (version 20.0, IBM Corp, Armonk, NY, USA).

## Results

28 patients were prescribed dupilumab for AD between January 2017 and August 2019. 14 patients were referred to the ophthalmology department with symptoms of bilateral eye redness, soreness, itching and epiphora with a mean time of onset of 6.75 (+/- 6.1) weeks from starting dupilumab. One patient was lost to follow up with the remaining 13 cases seen in the ocular surface clinic under the care of the ophthalmology department. There were no significant differences in demographic characteristics of patients treated with dupilumab who developed ocular symptoms and those who did not develop ocular symptoms.

Four patients seen in the ophthalmology department showed reduced tear break up time (TBUT <10 seconds) with meibomian gland disease and were diagnosed as having evaporative dry eyes without signs of conjunctivitis. They were prescribed lubricating eye drops and given advice on lid hygiene. One patient received a course of Maxitrol ointment for eyelid disease. They were excluded from further analysis.

Nine patients showed features of conjunctivitis on initial presentation with bilateral marked conjunctival redness [Figure 1]. Six patients displayed bilateral conjunctival papillary reaction and three had bilateral conjunctival follicular changes. Two patients showed limbal nodules similar in appearance to Trantas dots at initial visit.

**Figure 1.**
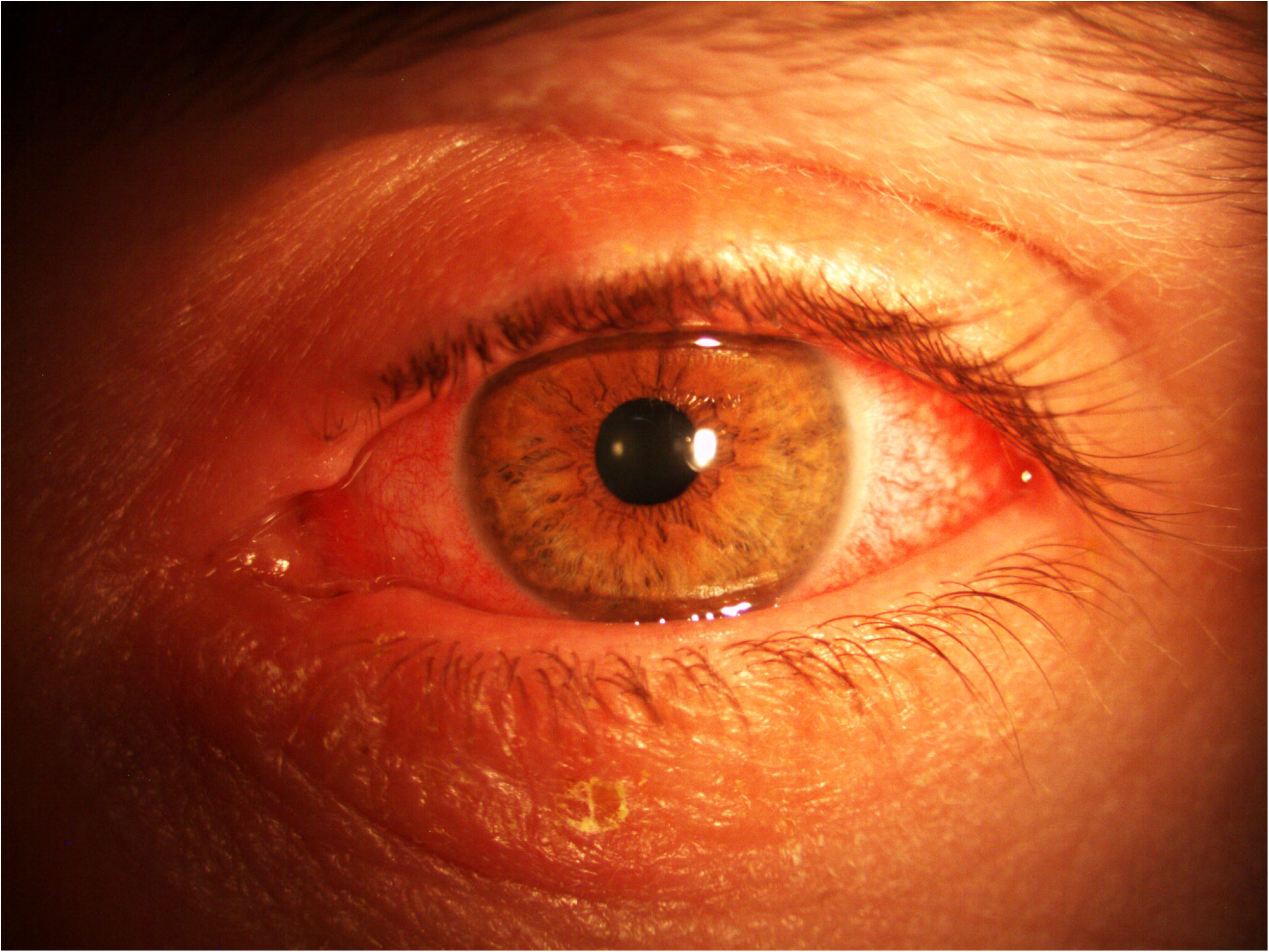
Patient with acute dupilumab - associated conjunctivitis.

Two female patients, who were related, had associated conjunctival cicatrisation in the lower fornix and one showed scarring in the superior tarsal conjunctiva on initial presentation. One of these patients went on to develop progressive cicatrisation and a leukoplakic lesion in the lower fornix, confirmed on biopsy as pre-cancerous actinic keratosis with dysplasia [Figure 2].

**Figure 2.**
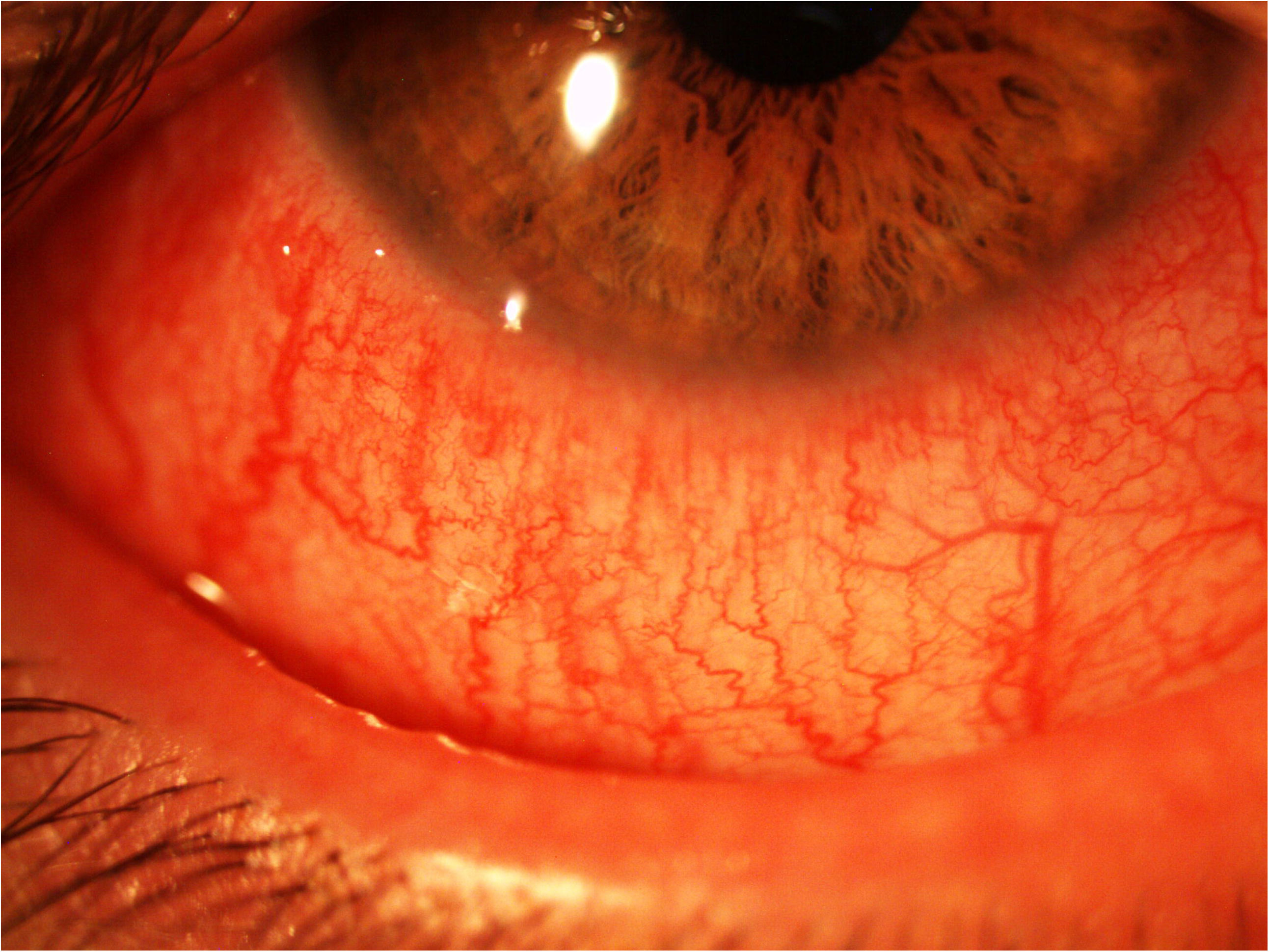
Patient on dupilumab with progressive cicatrisation and leukoplakic lesion in lower fornix.

Peri-ocular changes with cicatrising ectropion, punctal stenosis and periocular dermatitis were noted in four patients in association with conjunctivitis.

Mean time to development of self reported eye symptoms after the start of dupilumab therapy in the conjunctivitis group was 6 +/- 5.5 weeks. Patients were seen in the ocular surface clinic on average 8 +/- 5.2 weeks later. 44.4 % patients (4/9) had a past history of atopic keratoconjunctivitis prior to receiving dupilumab. Their ocular surface disease was stable at the time of dupilumab initiation with subsequent development of new symptoms of conjunctivitis. None were receiving active ophthalmic treatment at the time of starting dupilumab.

Initial treatment in the eye clinic for dupilumab-associated conjunctivitis included topical steroids in all cases. Six patients received additional topical lubricant. Out of nine patients presenting with conjunctivitis, seven had a good response to topical steroids alone [Figure 3], two required additional topical ciclosporin 0.1% (Ikervis®, Santen Pharmaceuticals) to control symptoms. One patient was prescribed tacrolimus ointment to the eyelids by the dermatologist.

**Figure 3.**
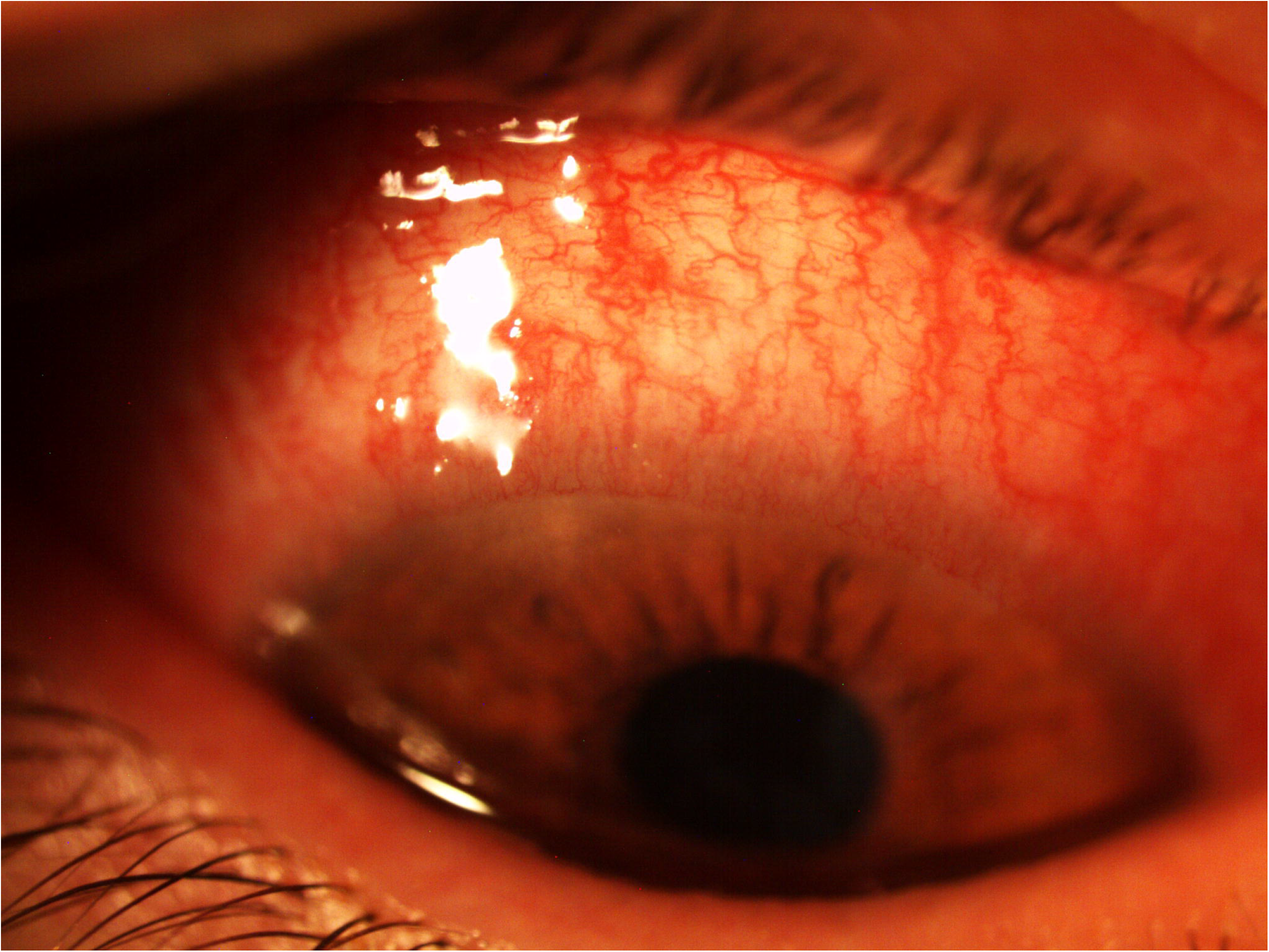
Patient from figure 1 after treatment with topical steroids.

67% (6/9) patients were on maintenance topical steroid or steroid and ciclosporin combination at a mean of 16 +/- 6.9 months follow up. Two of these six (33%) had a prior history of atopic keratoconjunctivitis, which was quiescent at the time of starting dupilumab. The remaining three patients used topical steroid for 3-12 months with complete resolution of symptoms and signs of conjunctivitis.

We have not noted any changes in visual acuity from first to last visit; mean visual acuity was recorded as 0.1 logMAR at both time points. No patients developed side effects due to prescribed drops. All patients had an improvement in their Eczema Area and Severity Index scores, with a 76% mean improvement in EASI score from pre-treatment to 16 weeks follow-up in the conjunctivitis group. 89% of patients achieved EASI 50 (50 % improvement in EASI score) and 55.6% achieved EASI 75 (75% improvement in EASI score).

One patient required additional therapy with methotrexate and one with topical steroids. Dupilumab was discontinued in one patient due to severe conjunctivitis with loss of eye-lashes (madarosis).

## Discussion

Conjunctivitis is a known side effect of dupilumab, particularly affecting AD patients.^1,2^ The LIBERTY AD CAFÉ study reported the highest incidence of dupilumab-associated conjunctivitis amongst initial trials (28%), attributed to higher awareness of this side effect amongst investigators.^1^ The CAFÉ trial reported conjunctivitis in 22.1% of patients, which was thought to be due to the high percentage of participants with severe AD and prior atopic keratoconjunctivitis, both of which are considered risk factors.^8,9^

In our series, we found a higher rate of dupilumab-associated conjunctivitis (32%) compared to landmark studies.^2^ This is in keeping with other clinical reports of real-world outcomes. Dupilumab-associated ocular surface disease was reported in 43% and 70% by Nahum et al. and Ivert et al. respectively^5,9^. These differences could be due to overall smaller numbers of patients included in the clinical reports compared to landmark studies.^2,4,5,10^

Maudinet et al ^10^ reported features of dupilumab-associated conjunctivitis in keeping with our findings. In their series, 60% of patients presented with mild injection and papillary conjunctivitis and were treated with topical lubricants, whereas the remaining 40% had severe follicular conjunctivitis requiring topical steroid treatment. In our cohort we did not find that differentiating between papillary and follicular presentation was clinically relevant. All patient had severe bulbar conjunctiva injection, associated more commonly with papillary response, and all necessitated steroid therapy.

Importantly, two patients in our study had cicatrising changes present in the lower fornix and subtarsal conjunctiva at presentation. These two patients were related, perhaps implicating a genetic predisposition for developing a more severe immune response to dupilumab. This could also be a result of their AD alone and / or due to the presence of chronic ocular inflammation. However, neither had a history of atopic keratoconjunctivitis (AKC) nor symptoms of chronic ocular surface problems requiring ophthalmic care prior to starting dupilumab. Another explanation for the severity of ocular surface inflammatory features in these two cases could be related to a delay in ophthalmic review between developing dupilumab related ocular symptoms and being seen in the ocular surface clinic. In one case this time delay was –five months.

One of the patients affected by cicatrisation went on to develop a progressive leukoplakic lesion in the lower conjunctival fornix, which was confirmed histologically to be actinic keratosis with dyplasia - a precancerous lesion. This was thought to be a result of life long immunosuppression and not solely a result of dupilumab use. This patient was continued on dupilumab due to good dermatological response. Further excisional biopsy with postoperative adjuvant treatment with Interferon alpha-2b drops was planned at last follow up.

One patient (3.5%) in our series had to discontinue dupilumab due to severe conjunctivitis and loss of lashes (madarosis). Madarosis has not been described before and represents an extreme reaction to dupilumab. ^2,5, 10-14^ To our knowledge, there are two other case reports of conjunctival cicatrisation associated with dupilumab use.^12,17^ Associated cicatrising ectropion, periocular dermatitis and punctal stenosis have been described more commonly making the term dupilumab-associated ocular surface disease rather than dupilumab-associated conjunctivitis more descriptive. ^10-15^

Dupilumab is also known to increase the risk of orofacial herpes simplex infection recurrence.^2,5^ Herpes simplex virus (HSV) uveitis while on dupilumab has been described with no reports of HSV keratitis.^5^ One patient in our series had previous history of HSV keratitis but did not develop any self-reported herpetic eye disease flare-ups whilst on dupilumab. No patients in our series were swabbed for the presence of HSV at the time of their acute presentation, so this cannot be ruled out for certain as a cause of their acute ‘dupilumab-associated’ conjunctivitis presentation. However, no patients demonstrated any of the typical features of herpes simplex keratitis and we consider HSV as the causative agent to be unlikely.

The aetiology of dupilumab-associated conjunctivitis is not fully understood but shares some common characteristics with atopic blepharoconjunctivitis and allergic eye disease - namely decrease in goblet cells, heightened OX40 ligand activity, eosinophilia and increased Demodex infestation due to changes in the ocular surface environment.^10,18-23^ Allergic blepharokeratoconjunctivitis is common in AD with 32-56% of patients affected.^18,22-24^ Previous history of atopic keratoconjunctivitis and more severe AD prior to dupilumab initiation have been implicated as risk factors for developing dupilumab-associated ocular surface disease.^8,9^ In our series, 44.4% of patients had prior history of allergic eye disease and 50% went on to require long-term ocular treatment.

All of our patients had improvement in ocular symptoms on topical steroids, which were the mainstay of the initial treatment.^25^ Topical steroids alone were sufficient at reversing signs and symptoms of acute conjunctivitis in 89% of patients. Two patients required additional ciclosporin drops to control their symptoms. Topical preparations of ciclosporin have been successfully used in treatment of ocular surface disease for many years. Ciclosporin is a calcineurin inhibitor that inhibits T-cell-mediated immune responses and has been shown to increase goblet cell numbers.^26^ Dupilumab targets IL-13 action and secondary depletion of goblet cells and mucin production in the conjunctiva could be responsible for dupilumab-associated conjunctivitis.^2-5^ Ciclosporin might be particularly useful in treatment of dupilumab-associated ocular surface disease as it could reverse some of the local immunological actions of dupilumab. Use of Ikervis (ciclosporin 0.1%) drops in dupilumab-associated ocular surface disease remains off label since Ikervis is approved for use in dry eyes.^28^ In our series, Ikervis drops used twice a day were added to topical steroids with good clinical outcomes. Use of tacrolimus^4,8^ and pimecrolimus^28^ ointments to the eye lids has also been reported to improve dupilumab-associated ocular surface disease in other case series..

Mean duration of follow up was 16 (+/- 6.9) months. A large proportion of our patients (67%) required continuation of topical immunosuppressive therapy to prevent flare-ups on discontinuation of drops. Akinlade et al. reported that 80% of patients had resolution of conjunctivitis while on dupilumab in clinical trials, but there was no mention of duration of symptoms, nor what therapy these patients received for treatment.^2^ It also leaves 20% of cases with on-going ocular surface inflammation while on dupilumab. These findings support our theory that dupilumab-associated conjunctivitis might not be just a one off event in all affected eyes. The drug might alter the ocular surface flaring up the susceptibility to chronic blepharoconjunctivitis in AD patients. Alternatively, a large percentage of AD patients considered for dupilumab have previously undiagnosed ocular surface inflammation. Clinically it is difficult to distinguish a blepharoconjunctivitis flare up from dupilumab-associated ocular surface disease as they share many similarities.^2,9-18^ In the series of Maudinet et al.,64% of patients had previously undiagnosed blepharoconjunctivitis when examined prior to starting dupilumab. Interestingly, lower rates of dupilumab-associated conjunctivitis were observed in patients seen and treated by the ophthalmologist prior to starting the drug than in patients not seen by ophthalmologist prior to dupilumab therapy (13% versus 25% respectively).

Current practice in the UK is that the ophthalmologist reviews patients after they develop ocular symptoms from dupilumab. Given that a large percentage of patients require long-term ocular treatments, dupilumab is considered in a group of patients with high risk for ocular surface problems, and dupilumab-associated conjunctivitis can result in cicatrisation, our recommendation is that AD patients should be examined by the ophthalmologist prior to starting the drug.

No patient in our cohort developed side effects due to topical therapy during the follow up and vision remained excellent. All had improvement in EASI scores. With the retrospective nature of our study and small sample it is impossible to say if our patients had pre-existing undiagnosed AKC or whether dupilumab sensitized their eyes to become chronically inflamed. Larger long-term prospective studies would be needed to answer this question. We also lack data on the exact duration of the acute dupilumab–associated conjunctivitis due to retrospective data collection. In our series of patients, those not requiring maintenance steroid drops used topical steroids between 3-12 months.

In summary, dupilumab is used in patients who have a strong predisposition to chronic ocular surface problems. In our series, 32% were affected by acute dupilumab-associated ocular surface disease, which responded well to topical steroids with or without adjunctive topical ciclosporin. Dupilumab-associated conjunctivitis can lead to conjunctival cicatrisation and be associated with periocular skin changes, punctal stenosis and madarosis, making dupilumab-associated ocular surface disease a more useful, catch-all term. Dupilumab may unmask a predisposition to chronic ocular inflammation and a significant number of patients need long-term topical immunosuppressive therapy. Patients should be examined by the ophthalmologist prior to starting dupilumab to optimise their ocular surface and arrange timely follow up to detect acute dupilumab-associated ocular surface disease more swift

## Data Availability

The data that support the findings of this study are available on request from the corresponding author. Data will be submitted to peer reviewed journal.

---

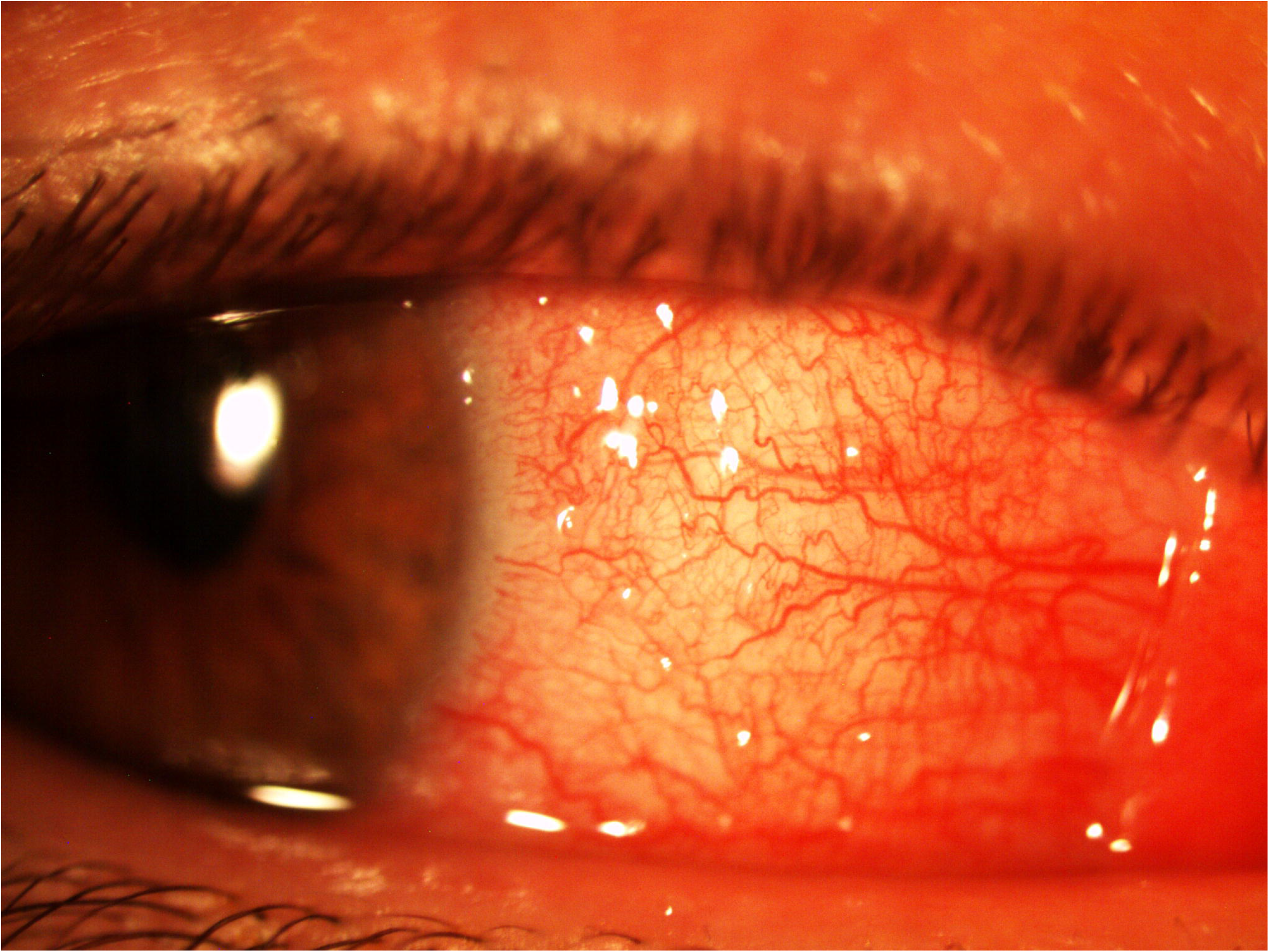

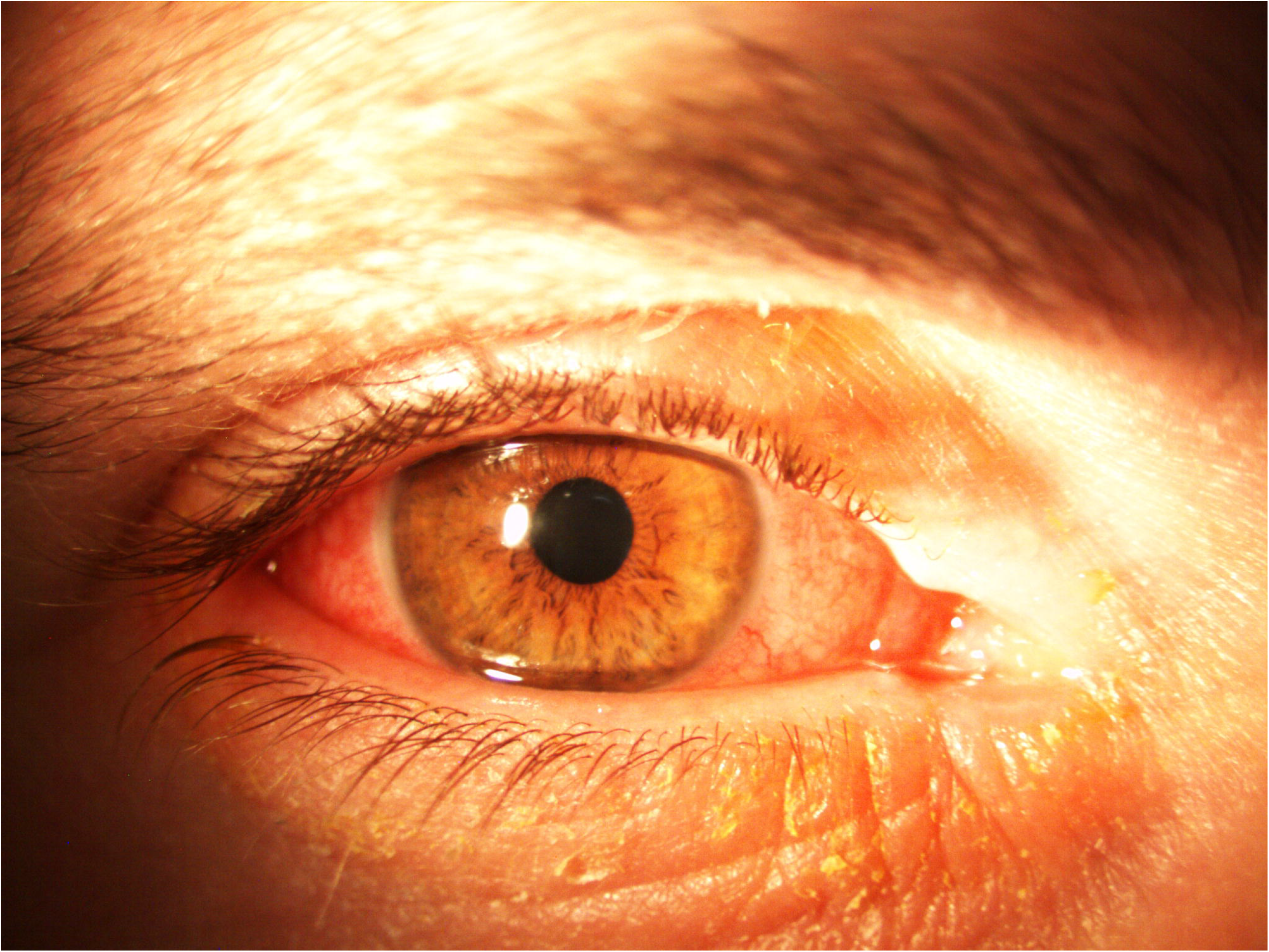

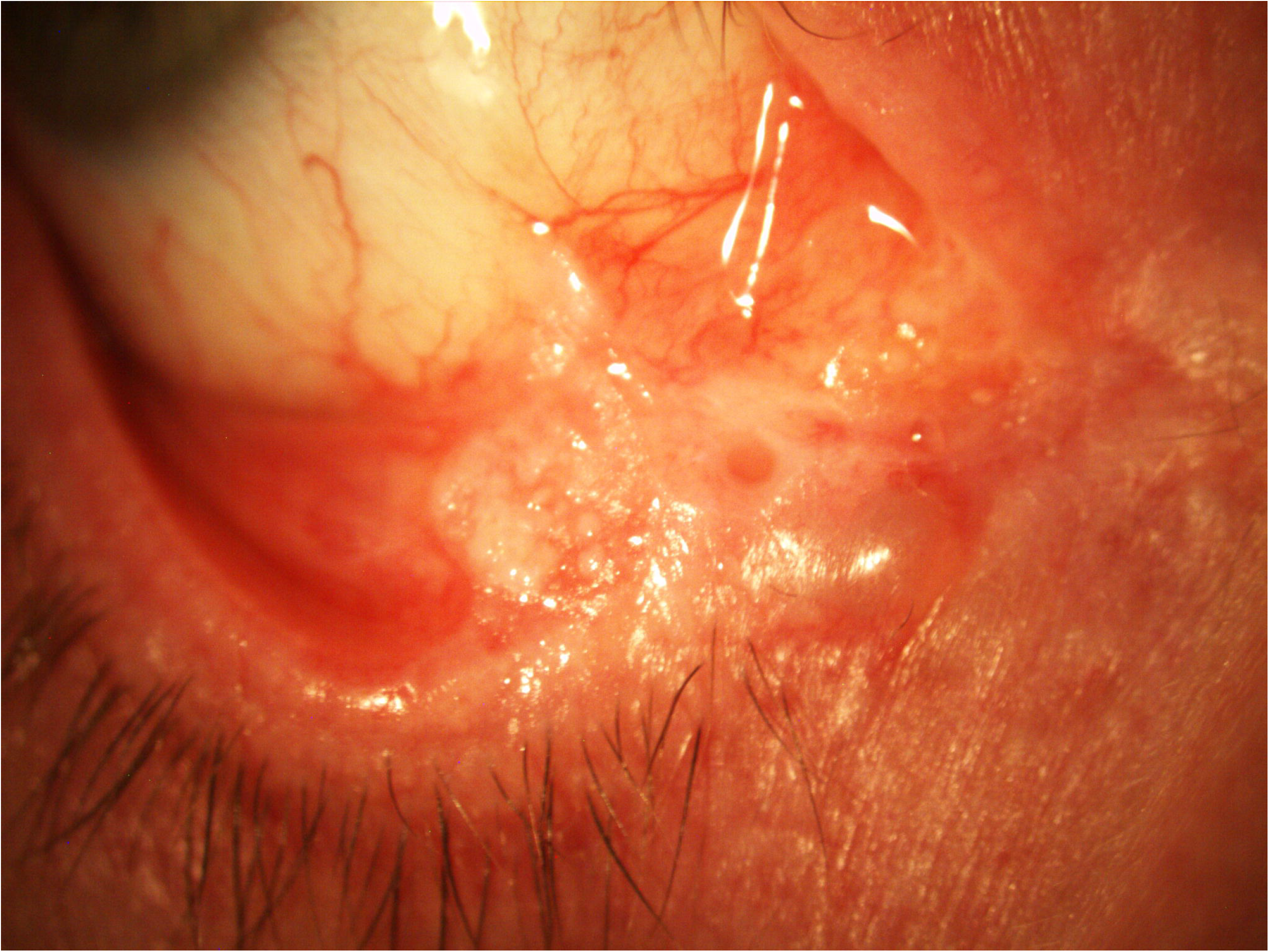

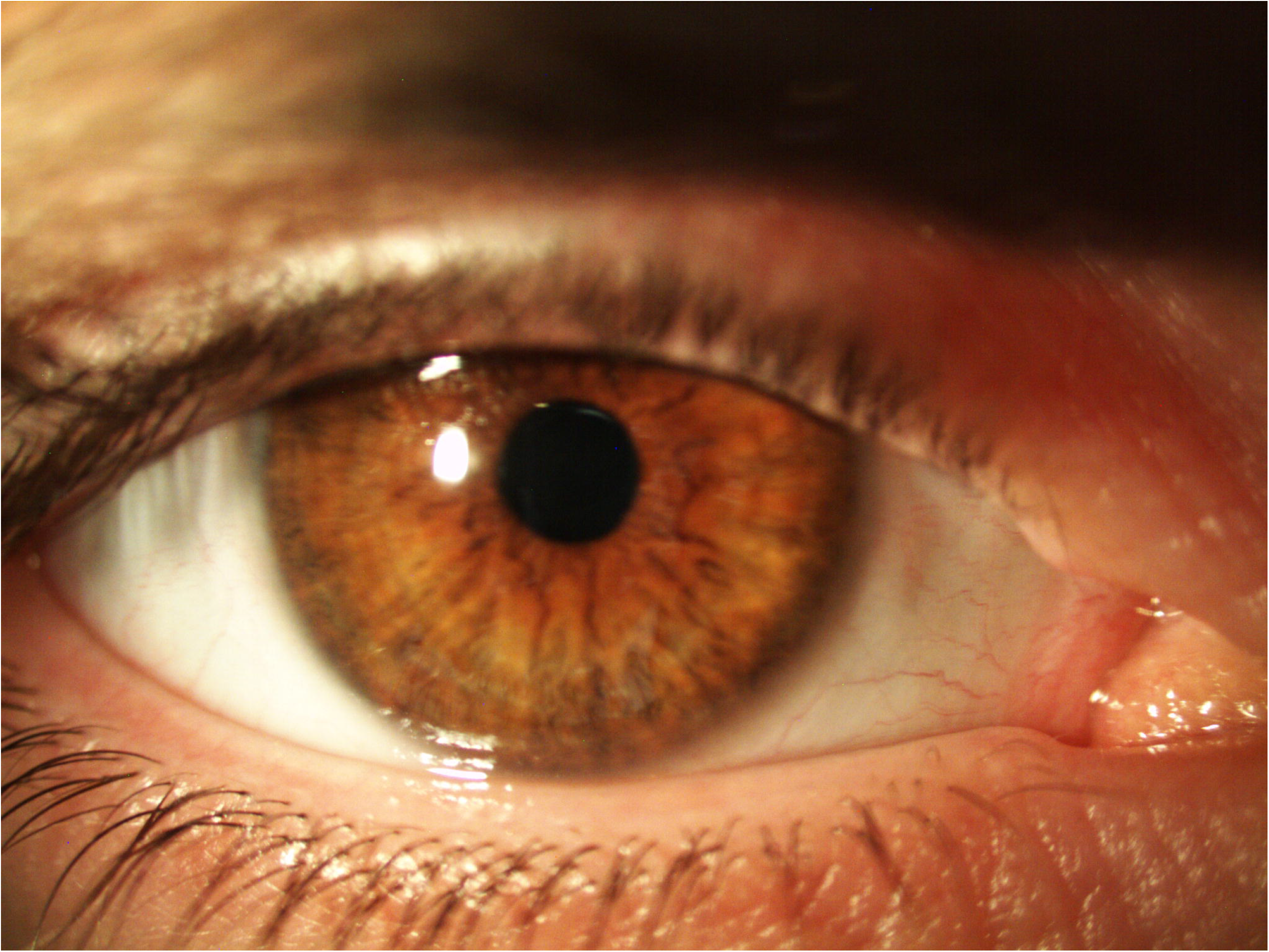

## Notes

### Competing Interest Statement

The authors have declared no competing interest.

### Funding Statement

No funding received for the study.

### Author Declarations

The Health Research Authority and Health and Care Research Wales (HCRW)

